# Imbalance-Aware Optimal Transport Learning for Cost-Effective Diabetic Retinopathy Screening

**DOI:** 10.64898/2026.04.16.26351035

**Authors:** Sulaiman Afolabi, Min Shi

## Abstract

**Background:** Diabetic Retinopathy (DR) is one of the leading cause of vision loss and blindness. AI models have been instrumental in providing an alternative solution to real-life medical treatment which are costly and sometimes not readily available in developing and underdeveloped nations. However, most of the existing AI models are developed with high-quality clinical images that makes it difficult to use such models in low-resource settings. For this reason, this research focus on bridging this gap by developing a low-resource, mobile-friendly, and deployable deep learning (DL) model for the detection of DR using an imbalance-aware optimal transport (OT) learning approach.

**Methods:** We trained our proposed framework with both high-quality hospital-grade images and low-resource smartphone-acquired images, and evaluated with the original test set from the smartphone domain. We also curated three levels of smartphone image-degradation quality and reported results from multiple experiments with bootstrapping. All model evaluations were assessed using the AUC, Sensitivity, and Specificity. Our results were compared with empirical risk minimization (ERM), Prototype OT, and Sinkhorn OT methods.

**Results:** We used four strong backbone architectures in the assessment. With our framework, Mobilevit-s achieved the best performance: an AUC of 87%, sensitivity of 89%, and specificity of 95%. Meanwhile, the statistical significance performance test (95% CI) shows that the AUC results are in the range of approximately 84% to 89%. For sensitivity, the range is 81% to 96%, and for specificity, 93% to 96%. This result indicated a performance increase of more than 3-5% compared to baseline methods.

**Conclusion:** Our framework shows promising results for low-resource DR screening, which has a potential to benefit less-advantaged groups and developing nations.

## 1 Introduction

DR is an advanced complication of diabetes and causes preventable vision impairment in the world [1].The damage caused by this disease can be drastically reduced if it is detected early and treated accordingly [2]. Retinal imaging is a major approach used in comprehensive eye examination that helps to mitigate DR. However, it often requires expensive and sophisticated medical devices that can only be found in hospitals and standard medical centers. These devices may not be available to rural patients in most parts of the world, especially in developing and underdeveloped nations. As a result, a large percentage of people who have limited access to the resources needed are marginalized for accurate and timely DR treatment. To bridge this gap, low-cost and reliable screening approaches are desired.

Artificial intelligence (AI) models have demonstrated sufficient capability in automating the screening of DR with low cost and the removal of marginalization issues [3][4][5]. However, the majority of the existing AI models are developed from high-quality hospital imaging created by highly sophisticated medical devices, which may not be suitable in a low-cost resource environment like the one this research targets. The model generalization issues, often referred to as domain shift, are a result of DL models’ tendency to fail under varying conditions, such as different imaging equipment, imaging modalities, and even demographic factors like race and ethnicity [6]. For example, fundus imaging exhibits domain shift in which models trained with hospital images reported significant performance degradation when deployed or tested with low-resource images, such as smartphone images, because of the variation in image quality, lesions, field of view, and noise levels [7][8]. As a result, AI models in clinical practice and diagnosis remains underutilized [9].

Several researchers have explored the use of AI for the diagnosis of DR with medical imaging, especially with publicly available data. For example, [10] classifies DR using a multi-attention residual refinement architecture on the popular EyePACS [11] dataset to achieve an improvement of 2-5% across the different backbones analyzed.developed an asymmetric bi-classifier discrepancy minimization system for DR screening by minimizing data set skewness because the authors ascertained that the general prediction of neural network models trained on a highly imbalanced dataset will be biased towards the majority class. The approach performs significantly better when compared with the ERM and three other state-of-the-art (SOTA) methods.developed an uncertainty-aware ordinal DL for cross-dataset DR grading, which is based on ordinal evidential loss with annealed regularization to encourage calibrated confidence of the model under domain shift. This method was evaluated on a multi-domain dataset including APTOS, Messidor-2 and a subset of the eyePACS fundus dataset and it achieves a meaningful competitive accuracy on the held-out test set in each cases. However, all the above approaches fail to address domain-shift in low-resource screening like smartphone fundus imaging.

An effective solution to remove the domain-shift problem in DL models, is to have a domain-wise and labeled target dataset that can be used to fine-tune existing AI models so that they can maintain their efficient performance, however such domain-wise dataset are scarce, poor quality, and often very limited in quantity, which cannot be used to train a reliable model for deployment in low-resource settings. To address these challenges, this work proposes an imbalanced-aware, class-conditional OT-based domain adaptation framework for DR screenings. It uses high-quality clinical images as the source domain and low-cost smartphone-acquired images as the target domain. Our approach aims for robust and semantically consistent cross-domain alignment with limited target supervision. This enables the development of lightweight, generalizable models for resource-constrained environments.

## 2 Methods

Since our objective is to develop a model that can generalize from hospital-grade clinical images to low-cost non-clinical images, we adopted a domain adaptation process known as OT learning [14]. OT is a principle method for aligning probability distribution and has been widely adopted to reduce features discrepancies across domains. It is an established mathematical concept used in machine learning to compare and manipulate different probability distributions in the feature space.

Inspired by this fundamental concept, we propose *OptimalFund* (Fig. 1), which utilizes the OT learning theorem in mitigating domain shift between the hospital DR images and smartphone images used in this research. Specifically, the framework aligns class-conditional feature distributions by minimizing Sinkhorn distance (i.e transport cost) [15] between corresponding classes across different domains. Thus, this alignment brings semantically similar classes together in the feature space, enabling the framework to learn variations across both domains. By minimizing transport cost across same-classes, the model learns discriminative representation and groupings for the five grading classes of DR. Our concept does not only align inter-domain classes together, but also solves the problem of imbalance distribution associated with low-cost DR dataset which is usually long-tailed. A bar chart showing the number of images in the different classes of the smartphone dataset is presented in Fig. 3.

**Fig. 1:**
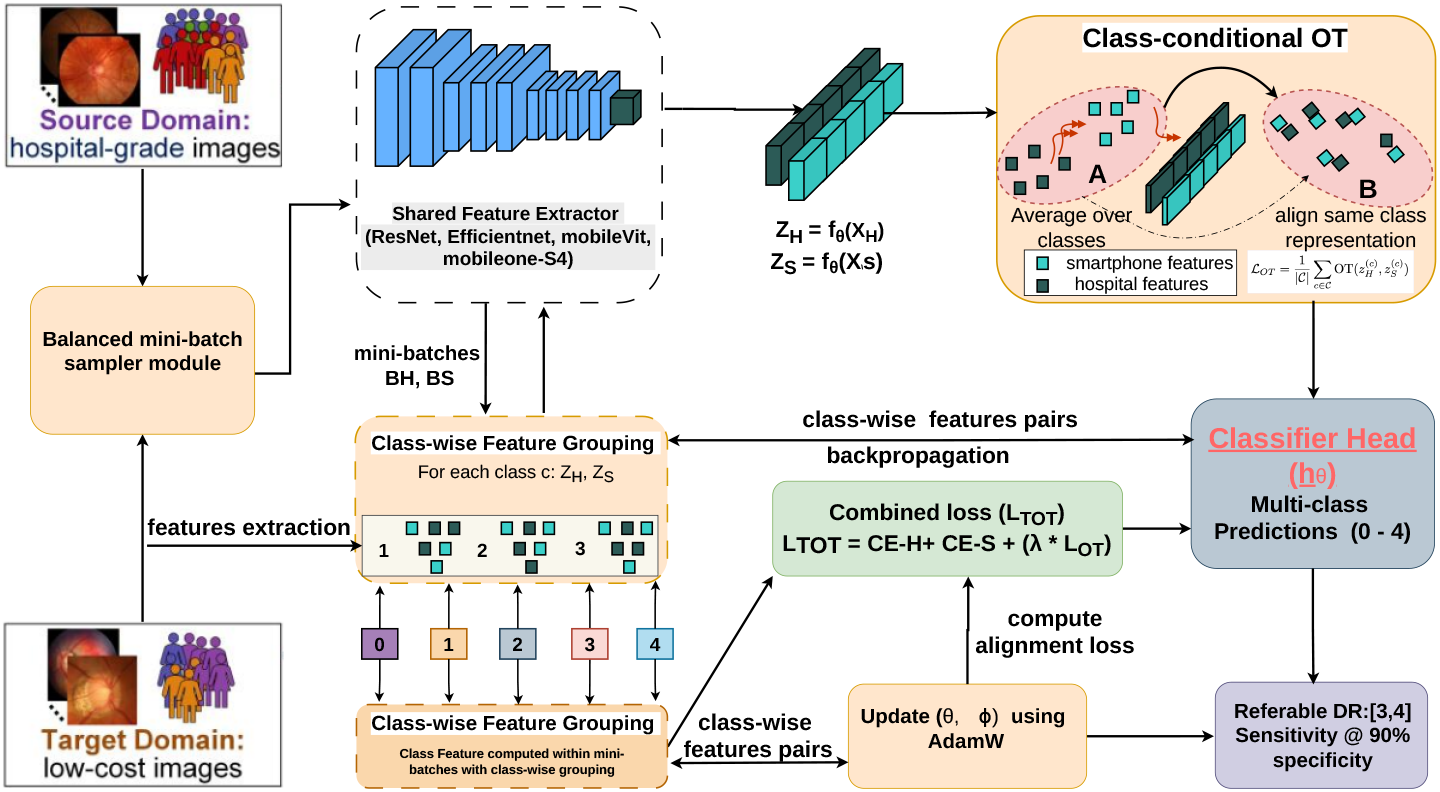
The Proposed OptimalFund Framework

### 2.1 Backbone Architecture

The proposed OptimalFund framework is integrated with different DL backbone architectures, with emphasies on the lightweight backbone architectures such as Efficientnet-b0, Mobileone-s4, and Mobilevit-s, which are very compatible with resource-constrained environments (devices). We compare the performance of our approach with ERM [16] and two other fundamental OT methods across multiple experimental settings and varying target-image dataset conditions that will be discussed in the subsequent part of this paper.

### 2.2 Dataset Description

In this study, two datasets were used to develop and evaluate DL models for the screening of multiclass DR. This study follows the declaration of Helsinki, in which all datasets were collected retrospectively, and the requirement for informed consent was waived. All patient personal information has been removed. The hospital DR dataset is used as provided by the data owner, as it is relatively balanced for usage, and already predefined into training, validation, and test sets. In contrast, the smartphone DR dataset required extra preprocessing. Specifically, we parsed the provided metadata to derive patient identifiers and labels for a stratified patient-level splitting in ratios of 0.7, 0.2, and 0.1 for the train, test, and validation sets, respectively. Both datasets DR labels consist of the standard five-level grading scale, which ranges from 0 - 4: (0 : No DR, 1 : mild NPDR, 2 : moderate NPDR, 3 : severe NPDR, 4 : proliferative DR). For clinical relevant evaluation metrics, sensitivity and specificity were computed using a binarized formulation, where classes 0-2 are classified as non-referable DR and classes 3-4 as referable DR. All images were loaded as RGB and resized to the acceptable model size(224 × 224), then standard augmentation techniques were applied for training purposes.

### 2.3 The Proposed OptimalFund Framework

The OptimalFund is a dual-level, model-agnostic framework specifically designed to handle domain-shift and class-imbalance, which can adversely affect the diagnostic performance of DL models. The core principle of this framework is to jointly learn feature representations from both domains while minimizing the disparity between class-conditional feature distributions in the shared feature space. Let *D*_*H*_ and *D*_*S*_ represent the hospital and smartphone datasets, respectively. Although, both domains share the same DR severity grading and a single encoder architecture, conventional training method will exhibit domain shift due to the difference in image quality, device, and acquisition style.

Our proposed framework consists of the shared feature extractor *f*_*θ*_ and the classifier head *h*_*θ*_ for multiclass prediction. The shared feature extractor is implemented as a DL backbone architecture which includes Resnet50 [17], Efficientnet-b0 [18], Mobilevit-s [19], and Mobileone-s4 [20]. It learns the feature representation from both domains simultaneously (Fig. 1). During training, mini-batches (equal sample size) from both domains are taken from the hospital and smartphone datasets via the balanced sampler before being fed to the Shared Feature Extractor (model). For each batch, the model extracts separate feature representations, and the supervised classification loss is computed across both domains. The supervised classification loss is used to learn discriminative DR features in both domains. To account for the severe class-imbalance in the smartphone domain, the framework has been integrated with an imbalance-aware sampling mechanism in the form of a class-balanced sampler. To further mitigate the domain gap, we introduce the class-conditional OT alignment module in the feature space. This modules align features distribution corresponding to the same DR class in both domains by minimizing the OT cost (distance) between their class distributions in the feature space. To preserve semantic representation across each class stages, we integrated a class-wise feature grouping module. This ensures that within each mini-batch, class-wise feature groupings are done to ensure corresponding feature alignment between the source and target domains. Class-wise features grouping prevents misalignment across the DR severity levels. The overall training objective is achieved by combining the supervised classification loss and the OT alignment loss, which is regulated by a hyperparameter *λ* to control the magnitude of the OT module. The OptimalFund framework is trained end-to-end with *AdamW* [21] as the optimizer. Mathematically,

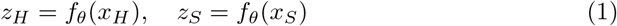

represents both features extracted from the hospital and smartphone mini-batches and

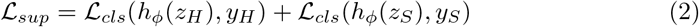

represents the classification loss on both batches where *L*_*sup*_ is the supervised classification, *L*_*cls*_ is the individual classification obtained from standard cross entropy (CE) loss [22]. For the OT alignment loss, it is given by the equation 3 below:

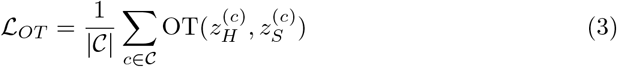

where C represents the number of classes in both domains. The overall training loss is presented in the equation 4 below:

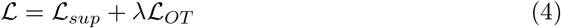

To evaluate OptimalFund, both hospital and smartphone datasets are jointly utilized during training. Specifically, training samples from both domains are fed into the framework simultaneously, while model validation is performed exclusively on the smartphone validation set. This is very important because the models need to prioritize the intended target domain (i.e smartphone images). We evaluated the framework on both standard backbone and lightweight architectures, which include Resnet50, Efficientnet-b0, Mobilevit-s, and Mobileone-s4. For each backbone, we selected the best model checkpoint across training epochs with the best validation AUC and the sensitivity at 90% specificity as the secondary criterion.

### 2.4 Smartphone Image-quality Degradation

To evaluate the robustness of the proposed OptimalFund framework, we introduce a controlled image quality degradation strategy on the smartphone datasets. This is to assess the performance of our framework under image quality worsening. We created three controlled degradation levels from the original datasets, which are the mild, medium, and severe(Fig. 2). For the mild level, we applied a Gaussian blur (radius = 0.8), additive Gaussian noise (*σ* = 6), JPEG compression (quality = 65), and reduced color(0.95), contrast(0.95), brightness(1.00), and vignette (strength = 0.10). The moderate level increases the severity of these transformations, which includes additive Gaussian blur (radius = 1.6), additive Gaussian noise (*σ* = 14), JPEG compression (quality = 40) and reduced color(0.85), contrast(0.85), brightness(0.98) and vignette (strength = 0.25). The severe level has the most aggressive degradation in the following order: additive Gaussian blur (radius = 2.8), additive Gaussian noise (*σ* = 26), JPEG compression (quality = 18), and reduced color(0.60), contrast(0.70), brightness(1.05), and vignette (strength = 0.55). These degradation settings are designed to simulate realistic artifacts common to smartphone-acquired fundus images. Such artifacts usually include reduced sharpness, brightness, uneven illumination, bad sensor noise, and poor image compression. This controlled setup enables systematic evaluation of the model robustness under rapidly challenging imaging conditions. Each degradation level is applied consistently across the test set for fair comparison across all methods explored in this study.

**Fig. 2:**
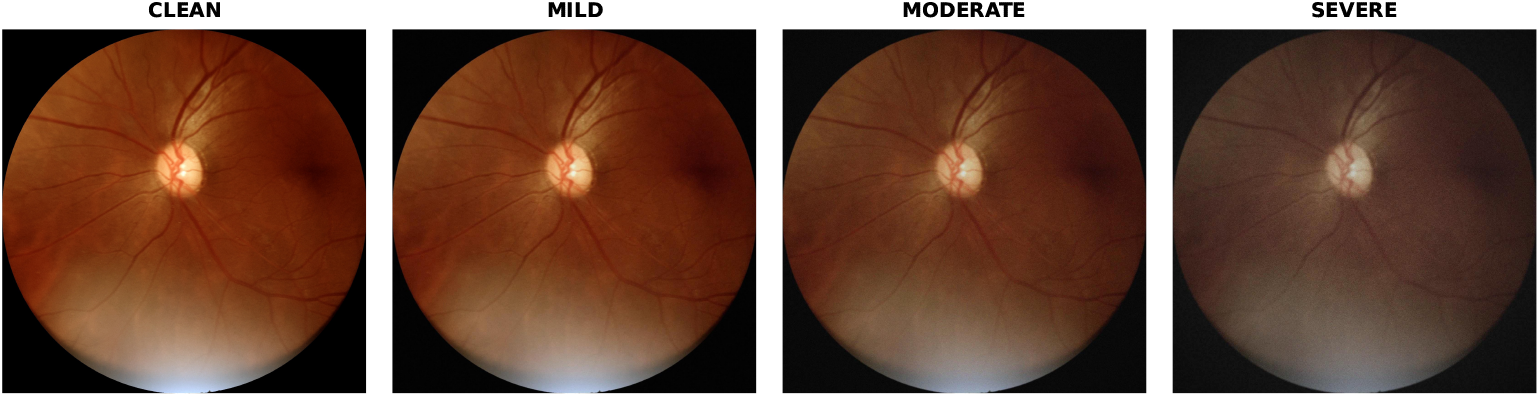
Smartphone Image-quality Degradation sample.

**Fig. 3:**
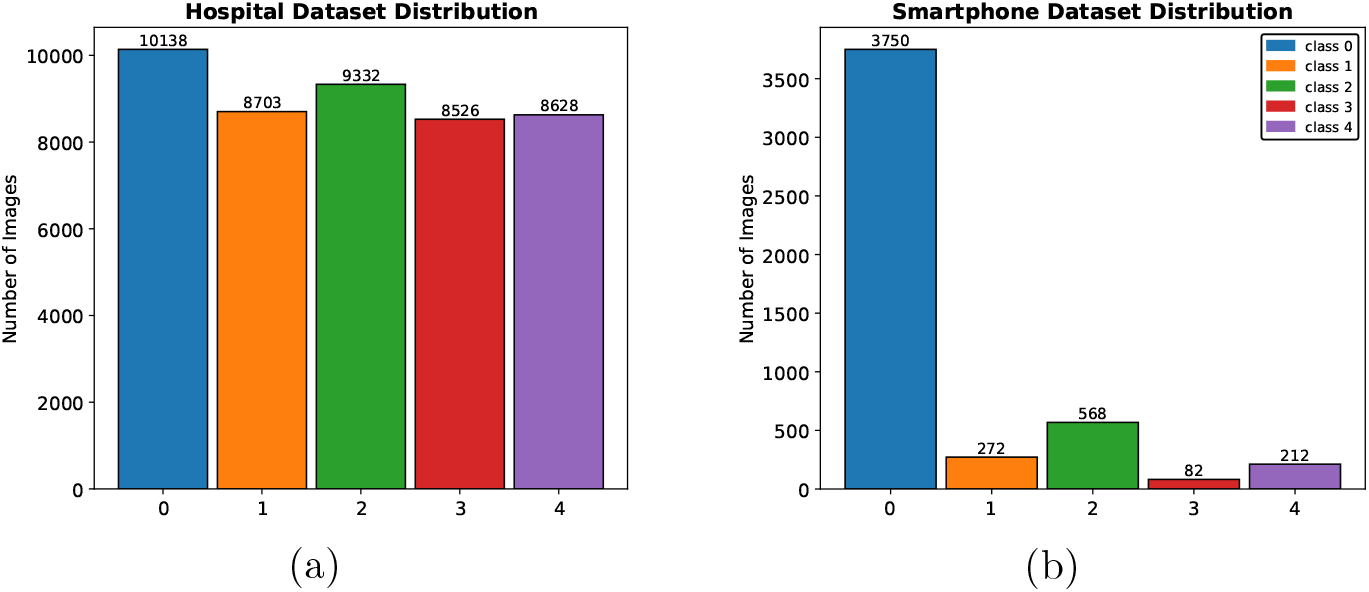
Class distribution comparison between the hospital and smartphone datasets.(a) Hospital dataset is relatively balanced. **b)** Smartphone dataset shows severe imbalance across the five DR classes.

### 2.5 Comparative method, Evaluation techniques and Statistical Analysis

We compare the proposed OptimalFund with ERM, prototype OT [23], and sinkhorn OT [24] baselines using the same training and evaluation protocol for DR screening. Experiments are conducted across four different DL architecture backbones notably for the medical image analysis, which include Resnet50, Efficientnet-b0, Mobilevit-s, and Mobileone-s4. We prioritize lightweight backbones in order to achieve a realistic deployment control and resource-constraint scenarios. All models performances are primarily assessed with the Area Under the Receivers Operating Characteristic Curve(AUC), along with sensitivity, and specificity to ensure clinical meaningful assessment, reliability, and efficiency. For each backbone, the best checkpoint was selected based on the smartphone validation AUC, and all reported test results were computed using the selected checkpoint. To ensure robustness, our statistical analysis focuses on the comparison among the OptimalFund, ERM, Prototype OT, and Sinkhorn OT results using multiple random seeds, and we reported the mean value results and standard deviation for experimental validity. In addition, bootstrap confidence interval are computed on the test set to estimate uncertainty in the models performance results.

### 2.6 Parameter and Implementation settings

All models were trained and evaluated with the same preprocessing and optimization settings to provide fair comparison and reporting integrity across all methods and backbones. All fundus images are loaded as RGB and resized to an input resolution of 224 × 224 for computational efficiency and speed optimization. Also, standard data augmentation such as random crop, horizontal flip, and color jitter (saturation, hue, brightness, contrast) is applied to the training dataset. In contrast, the validation and test sets are just resized and normalized for deterministic preprocessing. Model optimization is performed with *AdamW* optimizer with a fixed weight-decay of 1e-4. All model training is run for 30 epochs with a mini-batch size of 32 and a fixed multiple random seeds (42, 43, 44) for reproducibility purposes. For the proposed OptimalFund, we introduce additional parameters to regulate the effect of OT loss on the overall training loss. All model checkpoints were selected based on the best validation AUC on the smartphone dataset.

## 3 Results

### 3.1 Dataset Characteristics

As said earlier, we considered cross-domain datasets for the implementation of this idea. We used a pair of hospital images (high-quality clinical images with enhanced lesions) and smartphone image datasets (non-clinical images with low-cost acquisition and uneven illumination and field of view). The two datasets are a perfect example because of the difference in quality due to camera and imbalance distribution within the five classes. The datasets are available in public databases and collected for use in this research according to their respective licenses and data-use policies. The hospital domain dataset is a combination of DR fundus images curated from APTOS, DDR, EyePACS, IDRID, and Messidor, and it is obtained from *Kaggle*. Meanwhile, for the smartphone domain dataset, we make use of the Mobile Brazilian Retinal Dataset (mBRSET) collected using a smartphone with an hand-held portable camera. The smartphone dataset is obtained from the *Physionet* database [25] database after successfully acquiring the license. Both dataset domains follow the standard DR five-class (0-4) grading system, which consists of the NO DR, mild NPDR, moderate NPDR, severe NPDR, proliferative DR. The hospital DR dataset consists of 45327 images while the smartphone dataset consists of only 4884 images after adequate preprocessing and a stratified split at the patient level. The detailed overview of both domain dataset characteristics is presented in Table 1.

**Table 1:** Dataset summary for hospital and smartphone fundus images.

| Domain | Items | Statistics |
| --- | --- | --- |
| Hospital | Data type | Color fundus photographs |
|  | Label type | Multiclass (0: No DR, 1: Mild NPDR, 2: Moderate NPDR, 3: Severe NPDR, 4: PDR) |
|  | Class distribution | 0: 10,138 (22.4%), 1: 8,703 (19.2%), 2: 9,332 (20.6%), 3: 8,526 (18.8%), 4: 8,628 (19.0%) |
|  | Train / Val / Test | 29,840 / 6,796 / 8,691 (Total: 45,327) |
| Smartphone | Data type | Portable/smartphone fundus photographs |
|  | Label type | Multiclass (0: No DR, 1: Mild NPDR, 2: Moderate NPDR, 3: Severe NPDR, 4: PDR) |
|  | Class distribution | 0: 3,750 (76.8%), 1: 272 (5.6%), 2: 568 (11.6%), 3: 82 (1.7%), 4: 212 (4.3%) |
|  | Train / Val / Test | 3,420 / 493 / 971 (Total: 4,844) |

### 3.2 OptimalFund Results

Since we are focused on low-resource screening for DR, we treated the smartphone dataset as the target domain and we reported the multiclass AUC, referable clinical metrics (sensitivity at a fixed specificity of 90%) and the 95% bootstrapping confidence interval results using four different backbone models as shown in Table 2. The ERM AUC across the four backbone ranges from 80.73% to 84.36% with Mobilevit-s having the best performance, the prototype OT AUC ranges from 81.28% to 84.81%, the sinkhorn OT AUC ranges from 81.74% to 83.82%, while our OptimalFund AUC ranges from 81.69% to 86.53%, with notable improvement across all backbones. For example, comparing the ERM AUC result with our OptimalFund, Resnet50 improves from 81.52% to 81.69%, Efficientnet-b0 improves from 80.73% to 84.19%, Mobileone-s4 improves from 83.06% to 84.38% and Mobilevit-s improves from 84.36% to 86.53%.

**Table 2:**
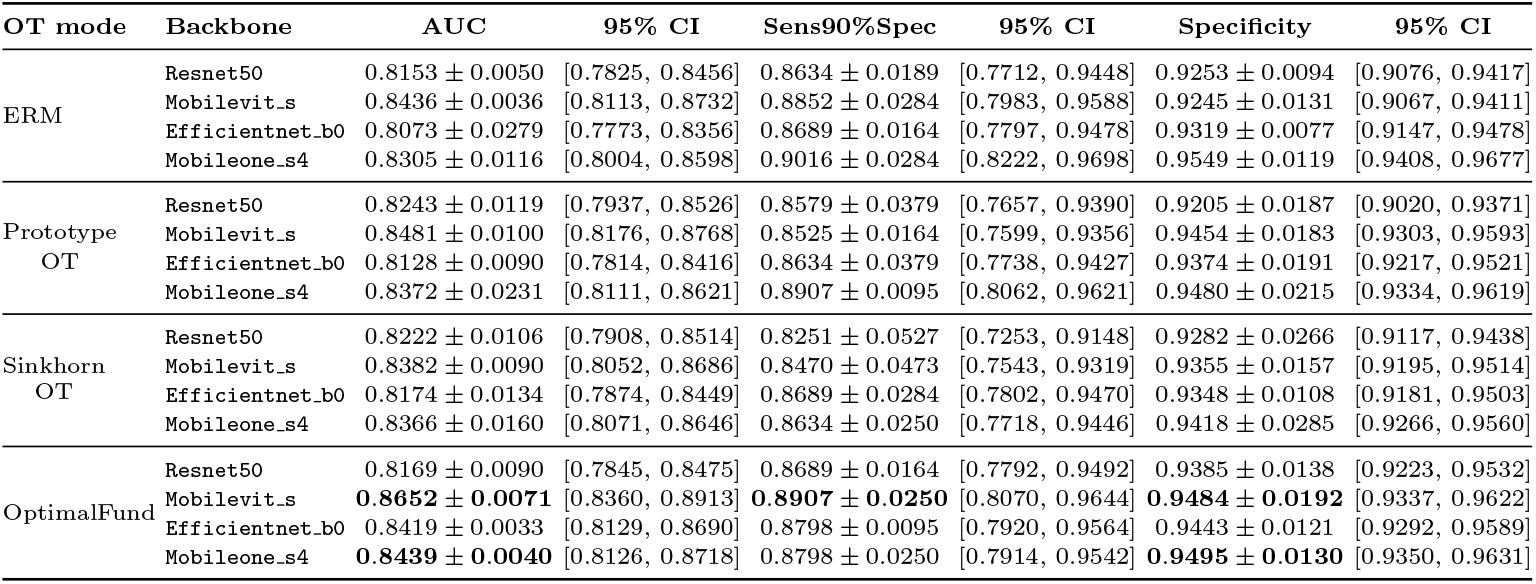
Test-set performance by OT mode and backbone for the original smartphone test split using multiple random seeds with 95% confidence interval bootstrap.

For sensitivity, the ERM values range from 86.33% to 90.16%, prototype values range from 85.24% to 89.07%, sinkhorn OT values range from 82.51% to 86.89% and the OptimalFund values range from 86.89% to 89.07%. For specificity, the ERM values range from 92.45% to 95.49%, prototype OT values range from 92.05% to 94.79%, sinkhorn OT values range from 92.82% to 94.17% while our OptimalFund specificity values range from 93.84% to 94.95%. These results further reinforced the clinical validity of our framework across all backbones considered in this research, as it demonstrated steady and reliable clinical metrics results.

### 3.3 Impact of Image Quality Degradation on DR Screening Results

Our OptimalFund framework shows consistent performance with different levels of smartphone image degradation when compared with the three other approaches evaluated in this research. For the mild degraded smartphone images results (Table 3), ERM AUC ranges from 79.69% to 83.44%, prototype OT AUC ranges from 80.44% to 84.31%, sinkhorn OT AUC ranges from 80.97% to 83.26% and OptimalFund AUC ranges from 80.81% to 84.38% which demonstrated robustness under poorer image condition. For the moderately degraded smartphone image results (Table 4), ERM AUC ranges from 79.22% to 82.02%, prototype OT AUC ranges from 80.07% to 83.40%, sinkhorn OT AUC ranges from 79.97% to 81.96%, while OptimalFund AUC ranges from 79.68% to 83.14% which is significantly better than the ERM results. For the severe version of the smartphone images results in Table 5, ERM AUC results ranges from 75.33% to 80.07%, prototype AUC ranges from 76.66% to 80.75%, sinkhorn OT AUC ranges from 76.30% to 79.91%, and OptimalFund AUC ranges from 78.14% to 80.42%.

**Table 3:** Test-set performance by OT mode and backbone for mild phone severity using multiple random seeds with 95% confidence interval bootstrap.

| OT mode | Backbone | AUC | 95% CI | Sens90%Spec | 95% CI | Specificity | 95% CI |
| --- | --- | --- | --- | --- | --- | --- | --- |
| ERM | Resnet50 | 0.8081 ± 0.0086 | [0.7760, 0.8381] | 0.8743 ± 0.0250 | [0.7858, 0.9503] | 0.8996 ± 0.0130 | [0.8793, 0.9186] |
| | Mobilevit.s | 0.8344 ± 0.0023 | [0.8031, 0.8622] | 0.9071 ± 0.0189 | [0.8279, 0.9759] | 0.8908 ± 0.0159 | [0.8697, 0.9106] |
| | Efficientnet.b0 | 0.7970 ± 0.0220 | [0.7668, 0.8258] | 0.8634 ± 0.0189 | [0.7730, 0.9435] | 0.9055 ± 0.0452 | [0.8869, 0.9234] |
| | Mobileone.s4 | 0.8145 ± 0.0046 | [0.7841, 0.8434] | 0.9126 ± 0.0189 | [0.8360, 0.9747] | 0.9114 ± 0.0374 | [0.8930, 0.9288] |
| Prototype OT | Resnet50 | 0.8179 ± 0.0036 | [0.7869, 0.8466] | 0.8525 ± 0.0434 | [0.7563, 0.9360] | 0.9004 ± 0.0380 | [0.8809, 0.9190] |
| | Mobilevit.s | 0.8431 ± 0.0109 | [0.8179, 0.8675] | 0.8962 ± 0.0250 | [0.8132, 0.9675] | 0.9139 ± 0.0308 | [0.8952, 0.9312] |
| | Efficientnet.b0 | 0.8044 ± 0.0119 | [0.7733, 0.8335] | 0.8634 ± 0.0413 | [0.7724, 0.9420] | 0.9234 ± 0.0181 | [0.9064, 0.9399] |
| | Mobileone.s4 | 0.8312 ± 0.0307 | [0.8055, 0.8563] | 0.9071 ± 0.0095 | [0.8271, 0.9736] | 0.9066 ± 0.0173 | [0.8869, 0.9253] |
| Sinkhorn OT | Resnet50 | 0.8097 ± 0.0134 | [0.7784, 0.8387] | 0.8525 ± 0.0434 | [0.7606, 0.9351] | 0.9015 ± 0.0345 | [0.8822, 0.9199] |
| | Mobilevit.s | 0.8326 ± 0.0038 | [0.8024, 0.8598] | 0.8907 ± 0.0189 | [0.8071, 0.9608] | 0.8978 ± 0.0211 | [0.8780, 0.9171] |
| | Efficientnet.b0 | 0.8100 ± 0.0142 | [0.7804, 0.8365] | 0.8852 ± 0.0164 | [0.8000, 0.9605] | 0.9055 ± 0.0154 | [0.8857, 0.9238] |
| | Mobileone.s4 | 0.8217 ± 0.0213 | [0.7920, 0.8499] | 0.8962 ± 0.0413 | [0.8152, 0.9651] | 0.9040 ± 0.0380 | [0.8852, 0.9225] |
| OptimalFund | Resnet50 | 0.8081 ± 0.0081 | [0.7760, 0.8382] | 0.8689 ± 0.0164 | [0.7771, 0.9498] | 0.9311 ± 0.0132 | [0.9145, 0.9472] |
| | Mobilevit.s | 0.8438 ± 0.0131 | [0.8151, 0.8699] | 0.8962 ± 0.0250 | [0.8141, 0.9668] | 0.9341 ± 0.0233 | [0.9177, 0.9497] |
| | Efficientnet.b0 | 0.8242 ± 0.0039 | [0.7962, 0.8510] | 0.8962 ± 0.0095 | [0.8138, 0.9666] | 0.9176 ± 0.0269 | [0.8998, 0.9350] |
| | Mobileone.s4 | 0.8224 ± 0.0059 | [0.7937, 0.8503] | 0.8962 ± 0.0250 | [0.8130, 0.9669] | 0.9227 ± 0.0358 | [0.9054, 0.9392] |

**Table 4:** Test-set performance by OT mode and backbone for moderate phone severity using multiple seeds and 95% confidence interval bootstrap.

| OT mode | Backbone | AUC | 95% CI | Sens90%Spec | 95% CI | Specificity | 95% CI |
| --- | --- | --- | --- | --- | --- | --- | --- |
| ERM | Resnet50 | 0.7922 ± 0.0160 | [0.7613, 0.8220] | 0.8689 ± 0.0328 | [0.7808, 0.9470] | 0.8604 ± 0.0211 | [0.8377, 0.8815] |
| | Mobilevit.s | 0.8202 ± 0.0021 | [0.7871, 0.8503] | 0.9016 ± 0.0328 | [0.8219, 0.9679] | 0.8289 ± 0.0604 | [0.8041, 0.8528] |
| | Efficientnet.b0 | 0.7955 ± 0.0284 | [0.7640, 0.8246] | 0.8853 ± 0.0164 | [0.8005, 0.9590] | 0.8865 ± 0.0503 | [0.8659, 0.9061] |
| | Mobileone.s4 | 0.8015 ± 0.0127 | [0.7696, 0.8319] | 0.9126 ± 0.0095 | [0.8361, 0.9781] | 0.8652 ± 0.0818 | [0.8440, 0.8860] |
| Prototype OT | Resnet50 | 0.8045 ± 0.0110 | [0.7735, 0.8330] | 0.8525 ± 0.0434 | [0.7553, 0.9355] | 0.8575 ± 0.0414 | [0.8347, 0.8785] |
| | Mobilevit.s | 0.8340 ± 0.0145 | [0.8074, 0.8597] | 0.9071 ± 0.0341 | [0.8285, 0.9729] | 0.8850 ± 0.0471 | [0.8644, 0.9046] |
| | Efficientnet.b0 | 0.8007 ± 0.0129 | [0.7686, 0.8300] | 0.8579 ± 0.0473 | [0.7667, 0.9394] | 0.9150 ± 0.0168 | [0.8965, 0.9330] |
| | Mobileone.s4 | 0.8246 ± 0.0270 | [0.7969, 0.8520] | 0.9180 ± 0.0164 | [0.8450, 0.9794] | 0.8725 ± 0.0198 | [0.8508, 0.8941] |
| Sinkhorn OT | Resnet50 | 0.8010 ± 0.0146 | [0.7699, 0.8308] | 0.8525 ± 0.0284 | [0.7587, 0.9374] | 0.8505 ± 0.0568 | [0.8278, 0.8727] |
| | Mobilevit.s | 0.8166 ± 0.0099 | [0.7868, 0.8452] | 0.8852 ± 0.0434 | [0.8019, 0.9574] | 0.8736 ± 0.0321 | [0.8519, 0.8948] |
| | Efficientnet.b0 | 0.7997 ± 0.0051 | [0.7689, 0.8280] | 0.8907 ± 0.0095 | [0.8065, 0.9650] | 0.8824 ± 0.0312 | [0.8614, 0.9020] |
| | Mobileone.s4 | 0.8196 ± 0.0232 | [0.7901, 0.8474] | 0.8962 ± 0.0189 | [0.8138, 0.9681] | 0.8795 ± 0.0422 | [0.8584, 0.9003] |
| OptimalFund | Resnet50 | 0.7968 ± 0.0069 | [0.7636, 0.8287] | 0.8743 ± 0.0189 | [0.7845, 0.9537] | 0.9209 ± 0.0227 | [0.9029, 0.9374] |
| | Mobilevit.s | 0.8314 ± 0.0112 | [0.8038, 0.8576] | 0.8798 ± 0.0250 | [0.7925, 0.9561] | 0.9330 ± 0.0333 | [0.9164, 0.9484] |
| | Efficientnet.b0 | 0.8154 ± 0.0005 | [0.7850, 0.8444] | 0.9071 ± 0.0095 | [0.8296, 0.9732] | 0.8985 ± 0.0324 | [0.8783, 0.9178] |
| | Mobileone.s4 | 0.8144 ± 0.0049 | [0.7853, 0.8424] | 0.8907 ± 0.0501 | [0.8073, 0.9646] | 0.8989 ± 0.0457 | [0.8793, 0.9178] |

**Table 5:**
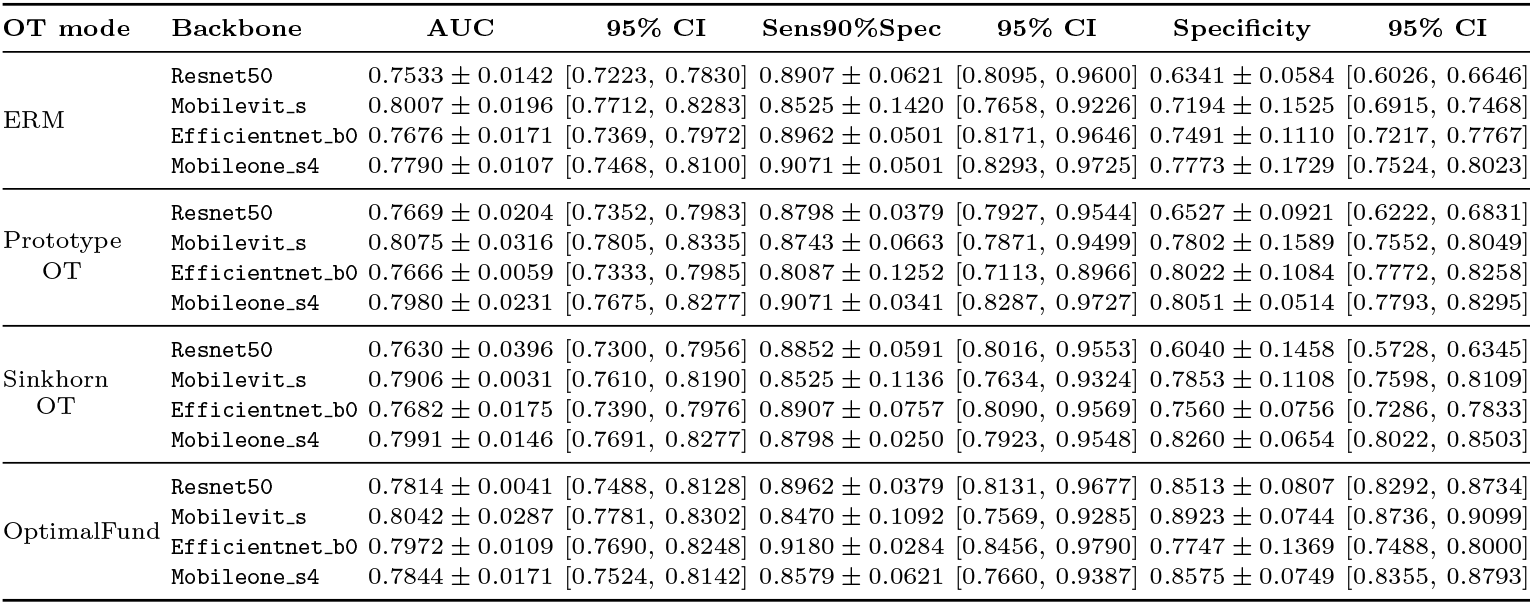
Test-set performance by OT mode and backbone for severe phone severity using multiple random seeds with 95% confidence interval bootstrap.

| OT mode | Backbone | AUC | 95% CI | Sens90%Spec | 95% CI | Specificity | 95% CI |
| --- | --- | --- | --- | --- | --- | --- | --- |
| ERM | Resnet50 | 0.7533 ± 0.0142 | [0.7223, 0.7830] | 0.8907 ± 0.0621 | [0.8095, 0.9600] | 0.6341 ± 0.0584 | [0.6026, 0.6646] |
| | Mobilevit.s | 0.8007 ± 0.0196 | [0.7712, 0.8283] | 0.8525 ± 0.1420 | [0.7658, 0.9226] | 0.7194 ± 0.1525 | [0.6915, 0.7468] |
| | Efficientnet_b0 | 0.7676 ± 0.0171 | [0.7369, 0.7972] | 0.8962 ± 0.0501 | [0.8171, 0.9646] | 0.7491 ± 0.1110 | [0.7217, 0.7767] |
| | Mobileone_s4 | 0.7790 ± 0.0107 | [0.7468, 0.8100] | 0.9071 ± 0.0501 | [0.8293, 0.9725] | 0.7773 ± 0.1729 | [0.7524, 0.8023] |
| Prototype OT | Resnet50 | 0.7669 ± 0.0204 | [0.7352, 0.7983] | 0.8798 ± 0.0379 | [0.7927, 0.9544] | 0.6527 ± 0.0921 | [0.6222, 0.6831] |
| | Mobilevit.s | 0.8075 ± 0.0316 | [0.7805, 0.8335] | 0.8743 ± 0.0663 | [0.7871, 0.9499] | 0.7802 ± 0.1589 | [0.7552, 0.8049] |
| | Efficientnet_b0 | 0.7666 ± 0.0059 | [0.7333, 0.7985] | 0.8087 ± 0.1252 | [0.7113, 0.8966] | 0.8022 ± 0.1084 | [0.7772, 0.8258] |
| | Mobileone_s4 | 0.7980 ± 0.0231 | [0.7675, 0.8277] | 0.9071 ± 0.0341 | [0.8287, 0.9727] | 0.8051 ± 0.0514 | [0.7793, 0.8295] |
| Sinkhorn OT | Resnet50 | 0.7630 ± 0.0396 | [0.7300, 0.7956] | 0.8852 ± 0.0591 | [0.8016, 0.9553] | 0.6040 ± 0.1458 | [0.5728, 0.6345] |
| | Mobilevit.s | 0.7906 ± 0.0031 | [0.7610, 0.8190] | 0.8525 ± 0.1136 | [0.7634, 0.9324] | 0.7853 ± 0.1108 | [0.7598, 0.8109] |
| | Efficientnet_b0 | 0.7682 ± 0.0175 | [0.7390, 0.7976] | 0.8907 ± 0.0757 | [0.8090, 0.9569] | 0.7560 ± 0.0756 | [0.7286, 0.7833] |
| | Mobileone_s4 | 0.7991 ± 0.0146 | [0.7691, 0.8277] | 0.8798 ± 0.0250 | [0.7923, 0.9548] | 0.8260 ± 0.0654 | [0.8022, 0.8503] |
| OptimalFund | Resnet50 | 0.7814 ± 0.0041 | [0.7488, 0.8128] | 0.8962 ± 0.0379 | [0.8131, 0.9677] | 0.8513 ± 0.0807 | [0.8292, 0.8734] |
| | Mobilevit.s | 0.8042 ± 0.0287 | [0.7781, 0.8302] | 0.8470 ± 0.1092 | [0.7569, 0.9285] | 0.8923 ± 0.0744 | [0.8736, 0.9099] |
| | Efficientnet_b0 | 0.7972 ± 0.0109 | [0.7690, 0.8248] | 0.9180 ± 0.0284 | [0.8456, 0.9790] | 0.7747 ± 0.1369 | [0.7488, 0.8000] |
| | Mobileone_s4 | 0.7844 ± 0.0171 | [0.7524, 0.8142] | 0.8579 ± 0.0621 | [0.7660, 0.9387] | 0.8575 ± 0.0749 | [0.8355, 0.8793] |

These results further reinforced that our OptimalFund shows consistent performance which is better than all other tested approaches, because it shows slower mean performance drops compared to others. With severe image quality degradation (Table 5), it is expected that all evaluated approaches will show significance performance drops compare to the other image-quality degradation results but the OptimalFund still maintain its superior stability across all backbones.

## 4 Discussion

There is no doubt about the rapid and continuous experimental research involving AI and DL in various fields, especially in the medical domain, however, its overall adoption remains short-lived [26]. With many excellent performance results recorded by AI systems, the overall adoption poses a challenge to question the sophisticated nature of these models, which is the focus of this research. Because most AI models with SOTA performance and quality are developed and evaluated using publicly available and clinically controlled datasets, it is challenging to evaluate such models in low-resource settings where they may fail to generalize. As a result, this study focuses on bridging the gap by developing a robust and generalizable AI model for DR screening, with a particular emphasis on low-resource settings, such as smartphone mobile deployment, to address the current challenges faced by less-advantaged groups in accessing and affording effective DR screening at low cost.

From the analysis of our results, OptimalFund maintains performance dominance over the other tested methods in this research. This claim is supported by various experimental results across different image quality degradation as shown in Fig. 4. By computing the average AUC performance of each OT mode across the different image-quality degradation levels, our proposed method show least performance drops across all backbones. Meanwhile, with sensitivity and specificity (Fig. 4), ERM shows the least performance drops in Average sensitivity but with worse specificity, while our OptimalFund demonstrated a clear and distinct average specificity across the image-quality degradation levels.

**Fig. 4:**
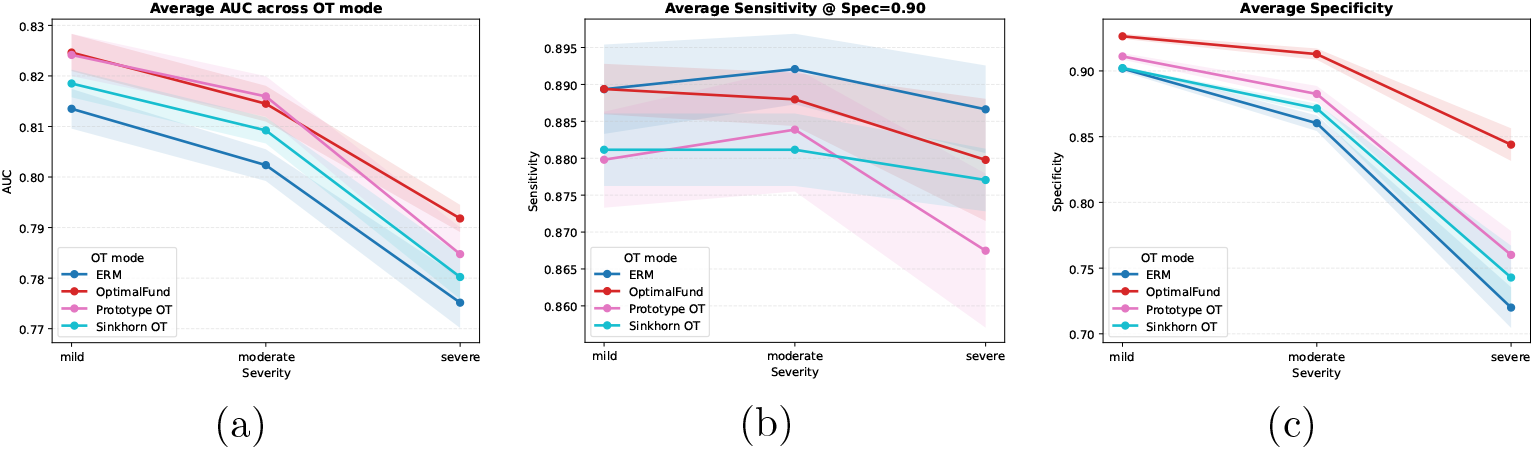
Performance degradation under increasing smartphone image severity across OT modes. **a)** AUC performance drop across OT modes. **b)** Sensitivity at 90% specificity. **c)** Specificity. Results are averaged across backbones.

Despite our method’s advantages, our study has some limitations. First, all experimental results and evaluations are based only on retrospective datasets. These datasets were sourced from various public domains that we do not control. Thus, we cannot guarantee it reflects and follows real-life medical data acquisition and guidelines. Second, we curated three image-quality degradation levels using our own settings. While this is useful for control experiments, we cannot confirm that our synthetic images cover all real fundus image artifacts. Third, the study only addresses multi-class DR classification using a few mobile-friendly backbones. In addition, it compares with several existing methods, but could be extended to include other ocular diseases, like glaucoma, and more backbone architectures. Finally, this study does not cover the deployment analysis or latency evaluation, which are important for real-world mobile deployment and implementation.

## 5 Conclusion

In conclusion, the most of the recent AI systems and DL models are developed and trained using controlled hospital (clinical) image datasets, which are usually incompatible for low-resource deployment and often fail when evaluated on non-clinical datasets (smartphone images). This issue has been attributed to poor domain generalization, termed domain shift. In this research work, we proposed an imbalance-aware OT learning-influenced DL framework that minimizes the transport cost between the source (hospital) domain and the imbalance-target domain (smartphone) in the feature space to assist model decision making for DR screening. We evaluated OptimalFund with ERM and two other approaches, and our approach demonstrated strong effectiveness for adequate DR classification. Future work will be to extend this framework to other ocular disease screening, especially when targeting low-resource screening.

## Data Availability

All data produced are available online at:
1. https://www.kaggle.com/datasets/sehastrajits/fundus-aptosddridirdeyepacsmessidor?select=split_dataset
2. https://physionet.org/content/mbrset/1.0/

